# Stage-dependent associations of Cardiovascular-Kidney-Metabolic Syndrome with Cognitive Decline, Depression, and Neuroimaging Signatures in multi-regional older adults

**DOI:** 10.64898/2026.10.21.26366180

**Authors:** Haoran Zhang, Yingqi Liao, Wanheng Zhang, Zhiying Lin, Darren M. Lipnicki, Hongwei Ji, Yifan Yan, Yaping Zhang, Henry Brodaty, Wei Wen, Nicole Kochan, Katya Numbers, Elena Rolandi, Sabrina Noel, Nikolaos Scarmeas, Dan Zhou, Xiao Tan, Perminder S. Sachdev, Tien Yin Wong, Kamen A. Tsvetanov, Xu Xin, the Cohort Studies of Memory in an International Consortium (COSMIC) group

## Abstract

**Background:** Cardiovascular–Kidney–Metabolic (CKM) syndrome is a multisystem condition associated with adverse outcomes, yet its impact on age-related cognitive health remains understudied. We examined associations of CKM with neurocognitive and neuropsychiatric outcomes, modifiable physical and psychosocial factors contributing to cognitive impairment across CKM stages, and neuroimaging correlates.

**Methods:** Global aging cohorts (mean follow-up 6.5 years) and the Cohort Studies of Memory in an International Consortium (5.4 years) were used. CKM stages (0–4) were defined according to AHA framework. Longitudinal domain-specific cognitive changes were assessed using linear mixed-effects models, and depressive symptoms using generalized linear regression with individual participant data approach. Population-attributable fractions were estimated for six modifiable factors associated with cognitive impairment across CKM stages. Cross-sectional associations with 34 neuroimaging biomarkers were examined in UK Biobank.

**Results:** Among 28,242 participants, advanced CKM stages (3–4) were associated with accelerated cognitive decline, particularly in attention, executive function, and language. Associations were stronger in younger individuals, men, and those with less education, but not modified by APOE ε4 status. Advanced CKM stages were associated with depressive symptoms among cognitively unimpaired individuals. Modifiable factors accounted for 24.8% of cognitive impairment risk across CKM stages, mainly social isolation (7.0%), depressive symptoms (5.2%), and physical inactivity (5.2%). Advanced CKM stages were linked to widespread MRI alterations (27 biomarkers), especially white matter hyperintensity.

**Conclusion:** Advanced CKM stages are associated with accelerated cognitive decline and greater cerebrovascular burden. Addressing psychosocial factors and physical inactivity, alongside management of CKM risk factors, may help reduce cognitive impairment.

## 1. Introduction

Cardiovascular–kidney–metabolic (CKM) syndrome is a recently established systemic disorder characterized by pathophysiological interactions among metabolic risk factors, chronic kidney disease (CKD), and cardiovascular system.^1^ Rather than considering these conditions in isolation, the 2026 AHA/ACC/ADA/ASN clinical practice guideline recommends defining CKM across five stages (i.e., CKM Stages 0-4) to capture progressive multisystem dysfunction, which helps support risk stratification and guide disease prevention across the disease continuum.^2^ Importantly, CKM is highly prevalent among older adults aged 65 years and older in both the United States and Asia and has been associated with adverse outcomes, such as mortality and cardiovascular disease.^3–5^ However, the association between CKM and brain health outcomes remains unclear, particularly across diverse ageing populations. Clarifying this relationship may help identify earlier neurocognitive and neuropsychiatric changes across CKM stages.

Emerging evidence has shown the associations between CKM stages and poorer cognitive outcomes, such as cognitive impairment and incident dementia.^6–9^ Biologically, the vascular and metabolic pathways driving CKM progression, including chronic inflammation, endothelial dysfunction, and cerebral small-vessel disease pathology, have been consistently linked to cognitive impairment and dementia.^10^ Notably, advanced CKM stages (3–4) are associated with a higher risk of incident dementia than earlier stages (0–2)^11^, suggesting a potential stage-dependent relationship between CKM severity and dementia risk. However, cognitive impairment may precede a clinical diagnosis of dementia by up to a decade^12^ and a critical research gap remains in understanding the long-term cognitive trajectories across the CKM stages preceding dementia onset. It also remains unclear whether demographic (e.g., age, sex, and education) and genetic (APOE ε4 carrier) factors may influence individual susceptibility to CKM-related cognitive decline. Furthermore, vascular and metabolic risk factors may differentially affect brain structure and function,^13,14^ , contributing to domain-specific cognitive decline that warrants further examination in preclinical stages.

Additionally, depressive symptoms may represent as an early neuropsychiatric manifestation associated with cognitive impairment and dementia^15^. Recent studies showed a significant relationship between increasing CKM stages and greater depressive symptoms,^5,16^ but this relationship remains less well characterized among cognitively unimpaired older adults. More broadly, depressive symptoms together with multiple modifiable lifestyle risk factors, such as physical inactivity and excessive alcohol consumption,^2,17^ may contribute differently to dementia risk across different CKM stages. Understanding their relative contribution to incident cognitive impairment across CKM stages may help more targeted strategies in early stages of the dementia continuum.

Leveraging large, population-based cohorts across diverse regions, this study aims to (1) characterize the associations of CKM syndrome with global and domain-specific cognitive decline longitudinally; (2) assess whether these associations varied according to age, sex, educational attainment, and APOE ε4 carrier status; and (3) investigate the association of CKM stages with depressive symptoms among cognitive unimpaired individuals, as well as evaluate the contribution of modifiable physical health and psychosocial factors to incident cognitive impairment across CKM stages. To further elucidate the potential brain structural changes underlying CKM, we identify potential neuroimaging biomarkers correlates of CKM syndrome. Analyses were conducted in global ageing cohorts and externally validated in an independent international consortium using an individual participant data (IPD) approach, with cross-sectional neuroimaging analyses performed in UK Biobank to provide exploratory insights.

## 2. Methods

### 2.1. Contributing studies and participants

This individual participant data (IPD) study utilized data from global aging cohorts,^18^ and validated in the data from Cohort Studies of Memory in an International Consortium (COSMIC).^19^

The global aging cohorts include the Survey of Health, Ageing and Retirement in Europe (SHARE),^20^ the English Longitudinal Study of Ageing (ELSA),^21^ and the China Health and Retirement Longitudinal Study (CHARLS),^22^ which were designed using harmonized survey protocols to enable cross-regional comparisons. Detailed descriptions of each study have been reported previously. The present study included data from SHARE waves 6–9 (2014–2022), ELSA waves 2–11 (2004-2024), and CHARLS waves 1–5 (2011–2020).

Four community-based longitudinal aging studies from COSMIC were included: Australia (MAS),^23^ Italy (Invece.Ab),^24^ Greece (HELIAD),^25^ and the United States (BPRHS).^26^ These cohorts varied in the number of waves (3–5) and follow-up duration (8–13 years), but all collected sufficient data to define CKM stages and assess both global and domain-specific cognition. Genotyping of the APOE gene (rs429358 and rs7412) was performed using polymerase chain reaction (PCR) amplification followed by restriction fragment length polymorphism (RFLP) analysis. Detailed information on study design, recruitment, inclusion/exclusion criteria, and data collection procedures for each cohort was provided in Supplemental Table 1.

To ensure comparability across diverse cohorts, systematic harmonization strategies were applied demographic, lifestyle, and clinical factors. Such harmonization is essential for reducing measurement heterogeneity. We excluded participants with dementia or memory diseases at baseline (Supplemental Table 2) and those lacking sufficient data to determine baseline CKM stage. Only participants with at least two cognitive assessments during follow-up were included. All contributing studies received ethical approval from relevant institutional committees, and study participants signed informed consent before the study. This study follows the Strengthening the Reporting of Observational Studies in Epidemiology (STROBE) guidelines (Supplemental Table 3). The study time frames, follow-up periods, and for each cohort are shown in Supplemental Figure 1.

UK Biobank (UKB) was used to conduct neuroimaging analyses, a large prospective cohort study including over 502,412 participants aged 37–72 years at baseline (2006–2010).^27^ Approximately four years after the initial assessment, follow-up imaging visits were conducted (from 2014 onward), during which brain magnetic resonance imaging (MRI) data were acquired for more than 40,000 participants.

### 2.2. Operational definition of CKM Staging

CKM stages were defined according to the AHA Presidential Advisory Statement on CKM Syndrome.^1^ This was as follows: (1) CKM stage 0 (No CKM risk factors): Normal BMI and waist circumference, and no criteria met for other stages; (2) CKM stage 1 (Excess or dysfunctional adiposity): Elevated BMI, elevated waist circumference, or prediabetes, without other metabolic risk factors or CKD; (3) CKM stage 2 (Metabolic risk factors and CKD): Presence of hypertriglyceridemia, hypertension, diabetes, CKD, or metabolic syndrome; (4) CKM stage 3 (Subclinical CVD in CKM): Subclinical CVD, severe kidney function loss, or kidney failure, with concurrent stage 1 or stage 2 features, or CKD; (5) CKM stage 4 (Clinical CVD in CKM): Clinical CVD or kidney failure, with concurrent stage 1, stage 2 features, or CKD. Components were assessed using blood biochemistry, self-reported health, and/or physical measurements, including body mass index (BMI), waist circumference, glycated hemoglobin A (HbA1c), fasting blood glucose (FBG), triglyceride, high density lipoprotein (HDL), total cholesterol, cystatin C (CysC), serum creatinine (Scr), hypertriglyceridemia, hypertension, diabetes, metabolic syndrome, CKD, subclinical CVD (defined as predicted 10-year CVD risk ≥20% with PREVENT equation),^28^ and clinical CVD. Operational definitions for each component within each cohort are provided in the Supplemental Table 4.

### 2.3. Cognitive outcomes

The primary outcome was cognitive decline. In the global aging cohorts, cognition was assessed using standardized tests, including immediate and delayed recall (adapted from Rey’s Auditory Verbal Learning Test), the Serial Sevens Test, and an orientation task (month, date, day, and year). For each cohort, raw scores from these tests were first summed and then standardized into z-scores using that cohort’s baseline mean and standard deviation (SD). A global cognitive z-score was subsequently derived as the mean of the cohort-specific standardized scores. This harmonization procedure was performed separately within each study before pooling.^29^ In COSMIC cohorts, cognition was evaluated through neuropsychological tests batteries administered at each wave. Raw scores were standardized into z-scores using baseline mean and SD values from participants without mild cognitive impairment or dementia in each cohort. Scores were pooled across waves, transformed to a Gaussian distribution while preserving percentile ranks, and winsorized at ±3 SDs. These were further normalized using a category-centered approach adjusted for age, sex, and education, with SDs derived from residuals of covariate-adjusted regression models.^30,31^ Tests were grouped into five domains—memory, attention, executive function, visuospatial ability, and language—based on established criteria (Supplemental Table 5). For domains assessed by multiple neuropsychological tests, a domain score was calculated as the standardized mean of the corresponding test scores. A global cognitive z-score was then derived as the standardized mean of the five domain scores. Incident cognitive impairment was implemented as the primary outcome in the global aging cohorts for analyses of multiple modifiable factors, enabling the estimation of relative risks and population attributable fractions (PAFs). Cognitive impairment was defined using a cutoff of a global cognitive z-score ≤ −1.5 standard deviations at any follow-up assessment.

### 2.4. Depressive symptoms and modifiable factors

In the global aging cohorts, depressive symptoms were assessed by European Depression (EURO-D) scale in SHARE and Centre for Epidemiologic Studies Depression Scale (CES-D) in CHARLS and ELSA. In the COSMIC studies, depressive symptoms were evaluated using CES-D in BPRHS and Geriatric Depression Scale (GDS-15) in HELIAD, Invece.Ab, and MAS. Depression scores were positively skewed with many zero values and were transformed using the inverse hyperbolic sine transformation.^32^ We further included modifiable factors that have been associated with dementia in the existing literature but are not incorporated into the definition of CKM stages. Six modifiable factors were selected: excessive alcohol consumption (present/absent), physical inactivity (present/absent), social isolation (present/absent), depressive symptoms (present/absent), hearing loss (present/absent), and vision loss (present/absent) (Detailed definitions in Supplemental Table 6). Both individual and overall PAFs were calculated to estimate the potential reduction in the risk of cognitive impairment if exposure to these modifiable factors were eliminated. Analyses of modifiable factors were conducted exclusively in the global aging cohorts, due to data availability and the large population-based sample size

### 2.5. Other covariates

Baseline questionnaires collected demographic and lifestyle information, including age, sex, socioeconomic status (SES), education levels (low, medium, and high). APOE genotype was only available in COSMIC cohorts and was categorized as having either at least one ε4 allele or none.

### 2.6. Neuroimaging biomarkers

MRI data were acquired using a Siemens Skyra 3T scanner (Siemens Healthcare, Erlangen, Germany) using a standard 32-channel head coil, according to a freely available protocol (http://www.fmrib.ox.ac.uk/ukbiobank/protocol/V4_23092014.pdf). As part of the scanning protocol, high-resolution T1-weighted images, three-dimensional T2-weighted fluid-attenuated inversion recovery (FLAIR) images, and diffusion data were obtained. High-resolution T1-weighted images were obtained using an MPRAGE sequence with the following parameters: TR = 2000 ms; TE = 2.01 ms; 208 sagittal slices; flip angle, 8°; FOV = 256 mm; matrix = 256 × 256; slice thickness = 1.0 mm (voxel size 1 × 1 × 1 mm); total scan time = 4 min 54 s. 3D FLAIR images were obtained with the following parameters: TR = 1800 ms; TE = 395.0 ms; 192 sagittal slices; FOV = 256 mm; 256 × 256; slice thickness = 1.05 mm (voxel size 1.05 × 1×1 mm); total scan time = 5 min 52 s. Diffusion acquisition comprised a spin-echo echo-planar sequence with 10 T2-weighted (b ≈ 0 s mm−2) baseline volumes, 50 b = 1000 s mm−2 and 50 b = 2000 s mm−2 diffusion-weighted volumes, with 100 distinct diffusion-encoding directions and 2 mm isotropic voxels; total scan time = 6 min 32 s. Structural imaging and diffusion data were processed by the UK Biobank team and made available to approved researchers as imaging-derived phenotypes (IDPs); the full details of the image processing and QC pipeline are available in an open-access article^27,33^.

This study included 34 neuroimaging biomarkers (Table S7) from T1-weighted and T2-weighted FLAIR structural neuroimaging, including brain structural and white matter tracts, the data were available in 29774 UK Biobank participants.

### 2.7. Statistical analysis

Participant characteristics were summarized as n (%) for categorical variables and mean ± SD for continuous variables, or median (interquartile range) for skewed distributions. Associations between CKM stage and cognitive decline were assessed using one-stage IPD analyses with linear mixed-effects models, including study and participant as random intercepts. Dependent variables were global and domain-specific cognition, with an interaction term for time in study (TIS) × CKM stage (treated as continuous and ordinal, with stage 0–1 or stage 0–2 as reference) as the primary exposure. TIS was calculated in years from baseline. A significant interaction term indicated an association between CKM stage and the annual rate of cognitive change. Covariates available in both the global aging and COSMIC cohorts were adjusted for, including age, sex, education, excessive alcohol consumption, depressive symptoms, and physical inactivity. To assess robustness, we also conducted a two-step IPD random-effects meta-analysis, pooling cohort-specific estimates from linear mixed-effects models to obtain main estimates of effect sizes. Between-study variance (τ²) was estimated using (Restricted maximum likelihood) REML. Heterogeneity and potential publication bias were evaluated using the I² statistic and funnel plots. Stratified analyses were conducted by age (>70 vs. ≤70 years), sex, education (high vs. low), and APOE ε4 status (carrier vs. non-carrier). Interactions were tested with likelihood ratio tests.

We examined the associations between CKM stages and depressive symptoms using generalized linear models among individuals without cognitive impairment, with study-specific estimates pooled using a two-step IPD random-effects meta-analysis. Then, associations between six modifiable factors and incident cognitive impairment were estimated using Cox proportional hazards models in the global aging cohorts, adjusting for the covariates described above. Follow-up started from the baseline assessment and continued until the onset of cognitive impairment, loss to follow-up, or the end of follow-up, whichever occurred first. Hazard ratios (HRs) and 95% CIs were estimated for each modifiable factor. Using the prevalence and HR of each factor, we calculated individual PAFs. These individual PAFs were then weighted by communalities—representing the proportion of shared variance among the factors— derived from a principal component analysis based on a tetrachoric correlation matrix. The weighted individual PAFs were subsequently combined to estimate the overall PAF. This approach has been applied in previous studies^17,34^. The associations between CKM stages and neuroimaging measures were assessed using multivariable linear regression models, with effect estimates expressed as fold changes. False discovery rate (FDR) correction was applied to account for multiple comparisons, and the fold changes of significant neuroimaging biomarkers were subsequently ranked and visualized.

Sensitivity analyses included: (1) assessing baseline differences between included and excluded participants; (2) using multiple imputation with the Markov Chain Monte Carlo method to address missing baseline covariates (10 imputed datasets); (3) restricting analyses to complete cases; and (4) excluding participants with possible undiagnosed dementia (diagnosed or self-reported memory disease at first follow-up). R Studio software version 4.1.2 was used, with a statistical significance level set at < 0.05.

## 3. Results

### 3.1. Sample characteristics

The study flowchart is presented in Supplemental Figure 2, and further details regarding exclusions in each study are provided in Supplemental Table 8. In the global aging cohorts, 24,883 participants with complete CKM data and at least one follow-up cognitive assessment were included, with a mean follow-up of 6.5 years (SD = 2.2). For the external COSMIC cohorts, 3,359 participants were included, with a mean follow-up of 5.4 years (SD = 3.0). Sample characteristics are summarized in Table 1. In both the global aging and COSMIC cohorts, the majority of participants were at CKM stages beyond stage 1, while stage 0 accounted for the smallest proportion. Higher CKM stages were associated with older age, male sex, and unhealthier lifestyle behaviors—except for alcohol consumption in the global aging cohorts and smoking in the COSMIC cohorts. Compared with the global aging cohorts, participants in the COSMIC cohorts were older, had a higher proportion of highly educated individuals, reported lower rates of current alcohol consumption and physical activity, and exhibited a higher prevalence of depressive symptoms. Comparisons of baseline characteristics between participants included and excluded from the main analyses are presented in Supplemental Table 9. In both cohorts, excluded participants were generally older, had a higher prevalence of depressive symptoms and a higher educational level, but reported lower rates of current alcohol consumption, physical activity, and current smoking. Additionally, a higher proportion of women were excluded in the global aging cohorts.

**Table 1.**
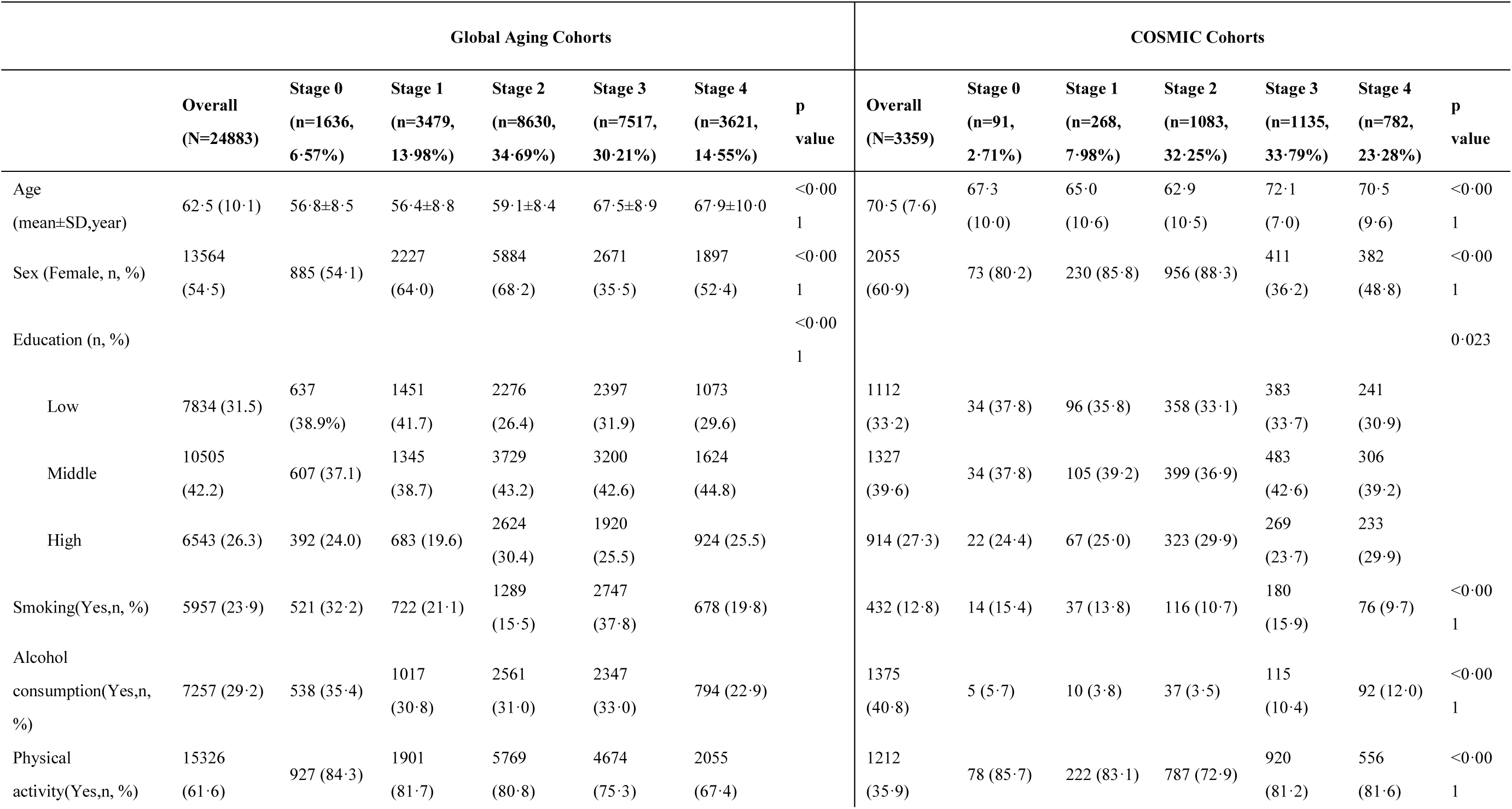
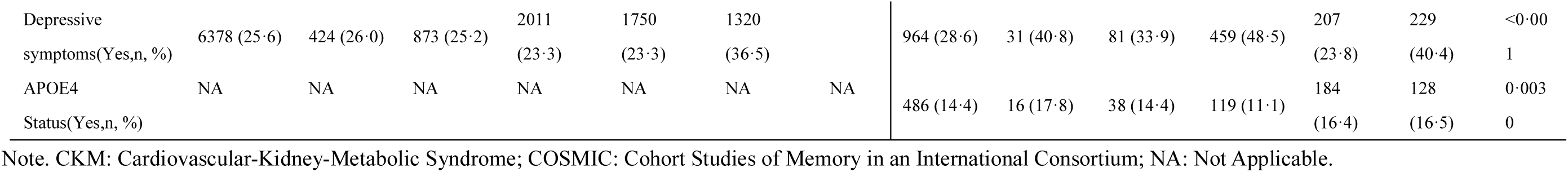
Baseline Characteristics by CKM stages of Study Participants.

### 3.2. Associations of CKM syndrome with global cognition

When treating CKM stages as a continuous variable in both global aging and COSMIC cohorts, greater CKM stages were consistently associated with a faster rate of decline in global cognitive performance (Table 2). When using CKM stages 0–1 as the reference, CKM stage 2 was not significantly associated with cognitive decline in either dataset. In the global aging cohorts, significant associations were observed at stage 3 (estimate = –0.031 SD/year, 95% CI: –0.037 to –0.026) and stage 4 (estimate = –0.031 SD/year, 95% CI: –0.038 to –0.025). Similar patterns were found in the COSMIC cohorts, with significantly faster declines in global cognition at stage 3 (estimate = –0.020 SD/year, 95% CI: –0.032 to –0.0077) and stage 4 (estimate = –0.022 SD/year, 95% CI: –0.043 to –0.016). Using CKM stages 0–2 as the reference, the global aging cohorts showed accelerated decline at stage 3 (estimate = –0.028 SD/year, 95% CI: –0.032 to –0.024) and stage 4 (estimate = –0.028 SD/year, 95% CI: –0.034 to –0.023). The COSMIC cohorts exhibited comparable associations, with faster declines at stage 3 (estimate = –0.016 SD/year, 95% CI: –0.025 to –0.0084) and stage 4 (estimate = –0.026 SD/year, 95% CI: –0.035 to –0.017). Overall, participants at advanced CKM stages (3–4) experienced a 24.6% to 274.7% greater rate of decline in global cognition compared with those at stages 0-1 or 0-2 (Table 2).

**Table 2.** Associations of CKM Syndrome With Longitudinal Global Cognition in Global Aging and COSMIC Cohorts.

|  | Global aging cohorts |  |  | COSMIC cohorts |  |  |
| --- | --- | --- | --- | --- | --- | --- |
|  | Decline rate (95% CI) | Estimate (95% CI) | Difference(%) | Decline rate (95% CI) | Estimate (95% CI) | Difference(%) |
| Continuous variable,<br>per 1 increase in CKM<br>stage | -0.027 (-0.029, -0.025) | -0.011 (-0.013, -0.010)*** | NA | -0.072 (-0.077, -0.068) | -0.009 (-0.014, -0.005)*** | NA |
| CKM reference 0-1 | -0.012 (-0.016, -0.008) | REF | NA | -0.058 (-0.070, -0.045) | REF | NA |
| CKM stage 2 | -0.016 (-0.019, -0.013) | -0.004 (-0.009, 0.001) | 33.3 | -0.063 (-0.070, -0.055) | -0.0050 (-0.020, 0.010) | 8.7 |
| CKM stage 3 | -0.043 (-0.046, -0.040) | -0.031 (-0.037, -0.025)*** | 256.8 | -0.077 (-0.084, -0.069) | -0.019 (-0.033, -0.004)* | 32.5 |
| CKM stage 4 | -0.043 (-0.048, -0.038) | -0.031 (-0.037, -0.024)*** | 256.8 | -0.088 (-0.098, -0.078) | -0.030 (-0.046, -0.014)*** | 52.2 |
| CKM reference 0-2 | -0.015 (-0.017, -0.012) | REF | NA | -0.062 (-0.068, -0.055) | REF | NA |
| CKM stage 3 | -0.043 (-0.046, -0.040) | -0.028 (-0.033, -0.024)*** | 186.7 | -0.077 (-0.084, -0.069) | -0.015 (-0.025, -0.005)*** | 26.1 |
| CKM stage 4 | -0.043 (-0.047, -0.038) | -0.028 (-0.033, -0.022)*** | 186.7 | -0.088 (-0.098, -0.078) | -0.026 (-0.038, -0.015)*** | 45.8 |

Note. \*p < 0.05; \*\*p < 0.01; \*\*\*p < 0.001. CKM: Cardiovascular-Kidney-Metabolic Syndrome; COSMIC: Cohort Studies of Memory in an International Consortium; NA: Not Applicable. One step IPD analysis with linear mixed-effects models were used with adjustment for age, sex, educational attainment, drinking status, depressive symptoms, and physical activity. Decline rate represents estimated rate of annual change rate. Estimate describes the change in cognitive function (global z-scores) per year.

We further conducted two-step IPD analyses using random-effects models to validate these findings and observed consistent results (Figure 1). For example, CKM stage 2 was not significantly associated with longitudinal changes in global cognition in either the global aging or COSMIC cohorts. In contrast, advanced CKM stages (3/4) were associated with detrimental effects on cognitive decline, although the association for CKM stage 3 in the COSMIC cohorts (reference = CKM stages 0–2) did not reach statistical significance. Funnel plots are presented in Supplemental Figure 3.

**Figure 1.**
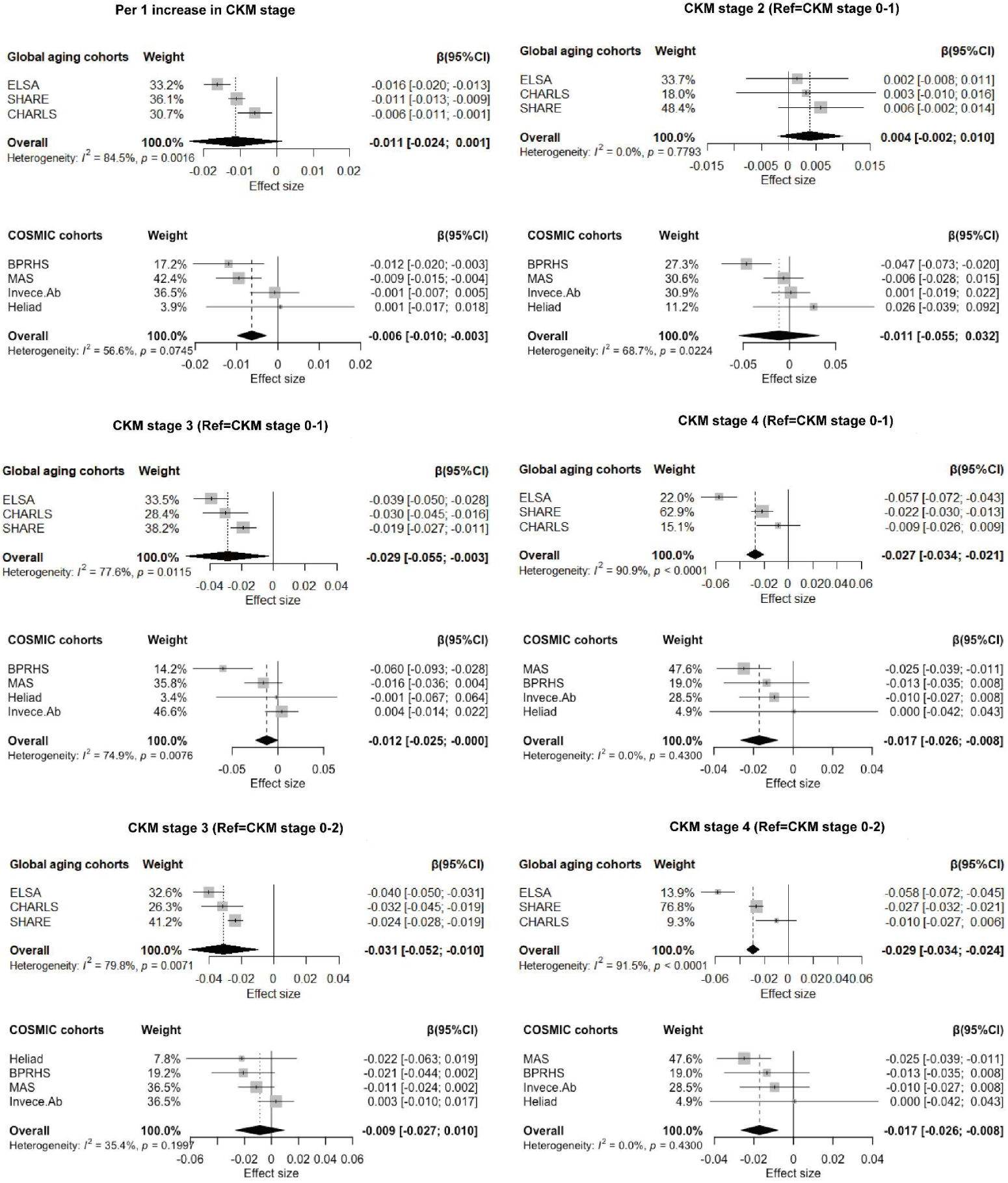
Forest plots from the 2-step IPD meta-analysis of the Associations of CKM Syndrome with Longitudinal Global Cognition in Global Aging and COSMIC Cohorts. Note. Two-step IPD meta-analysis using random effect models were conducted with adjustment for age, sex, education, excessive alcohol consumption, depressive symptoms, and physical inactivity. Estimate describes the change in cognitive function (global z-scores) per year.

We next conducted stratified analyses to assess whether the associations between CKM stages and cognitive decline varied across subgroups (Supplemental Figure 4). In the global aging cohorts, the adverse impact of advanced CKM stages on cognitive trajectories was more pronounced among participants aged ≤ 70 years (P for trend <0.05), male, and those with lower educational levels. In the COSMIC cohorts, stronger associations were consistently observed among younger participants and male (P for trend <0.05). APOE ε4 carrier status (evaluated only in COSMIC studies) did not modify the associations.

### 3.3. Associations of CKM syndrome with domain-specific cognition

The associations of CKM syndrome with domain-specific cognition were evaluated in the COSMIC cohorts, using CKM stage as either a continuous variable or categorical variables with stages 0–2 as the reference group (Table 3). We showed that each one-stage increase in CKM severity was significantly associated with a faster rate of decline in language (β = –0.0091 SD/year, 95% CI: –0.013 to –0.0052), attention (β = –0.0087 SD/year, 95% CI: –0.013 to –0.0042), and executive function (β = –0.0059 SD/year, 95% CI: –0.010 to –0.0016), with Bonferroni correction for multiple testing applied (adjusted significant threshold, p<0.003). These associations were also evident among individuals with advanced CKM stages (3/4) compared with those at stages 0–2 (Table 3). By contrast, increasing CKM stages were not significantly associated with memory or visuospatial speed (p > 0.05).

**Table 3.** Associations of CKM Syndrome With Longitudinal Domain-specific Cognition in COSMIC studies.

|  | Memory | Visuospatial | Language | Attention | Executive function |
| --- | --- | --- | --- | --- | --- |
|  | Estimate (95% CI) | Estimate (95% CI) | Estimate (95% CI) | Estimate (95% CI) | Estimate (95% CI) |
| Continuous variable, per 1 increase in CKM stage | -0.004 (-0.009, 0.001) | -0.006 (-0.011, -0.0006) | -0.009 (-0.013, -0.005)* | -0.009 (-0.013, -0.004)* | -0.006 (-0.010, -0.002)* |
| Reference = CKM stage 0~1 |  |  |  |  |  |
| CKM stage2 | 0.006 (-0.010, 0.022) | -0.010 (-0.028, 0.006) | 0.004 (-0.009, 0.018) | 0.005 (-0.010, 0.021) | 0.001 (-0.014, 0.016) |
| CKM stage3 | 0.004 (-0.012, 0.020) | -0.016 (-0.034, 0.002) | -0.017 (-0.030, -0.003)* | -0.017 (-0.029, -0.001)* | -0.016 (-0.031, -0.001)* |
| CKM stage4 | -0.012 (-0.029, 0.004) | -0.023 (-0.042, -0.004) | -0.022 (-0.036, -0.007)* | -0.021 (-0.038, -0.005)* | -0.015 (-0.030, -0.001)* |
| Reference = CKM stage 0~2 |  |  |  |  |  |
| CKM stage3 | 0.001 (-0.010, 0.011) | -0.006 (-0.018, 0.005) | -0.020 (-0.029, -0.011)* | -0.018 (-0.028, -0.008)* | -0.016 (-0.026, -0.007)* |
| CKM stage4 | -0.015 (-0.027, -0.003) | -0.014 (-0.027, -0.001) | -0.025 (-0.036, -0.015)* | -0.025 (-0.037, -0.014)* | -0.015 (-0.026, -0.004)* |

Note. \*p < 0.05 where p was adjusted using Bonferroni correction. CKM: Cardiovascular-Kidney-Metabolic Syndrome; COSMIC: Cohort Studies of Memory in an International Consortium. One step IPD analysis with linear mixed-effects models were used with adjustment for age, sex, educational attainment, drinking status, depressive symptoms, and physical activity. Estimate describes the change in cognitive function (domain-specific z-scores) per year.

### 3.4. Associations of CKM stages with depressive symptoms and modifiable factors of incident cognitive impairment across CKM stages

The results further showed that advanced CKM, especially stage 4, was consistently associated with depressive symptoms among cognitively unimpaired individuals across global aging and COSMIC studies (Supplementary Table 10). All six modifiable factors were significantly associated with incident cognitive impairment (Supplemental Table 11). The overall weighted PAF for the six modifiable factors was estimated at 24.7% in the total sample, 24.4% among participants at CKM stages 0–2, and 24.8% among those at CKM stages 3–4 (Figure 2). At CKM stage 3–4, social isolation (7.0%), untreated vision loss (5.2%), and depressive symptoms (5.2%) accounted for the largest weighted PAFs.

**Figure 2.**
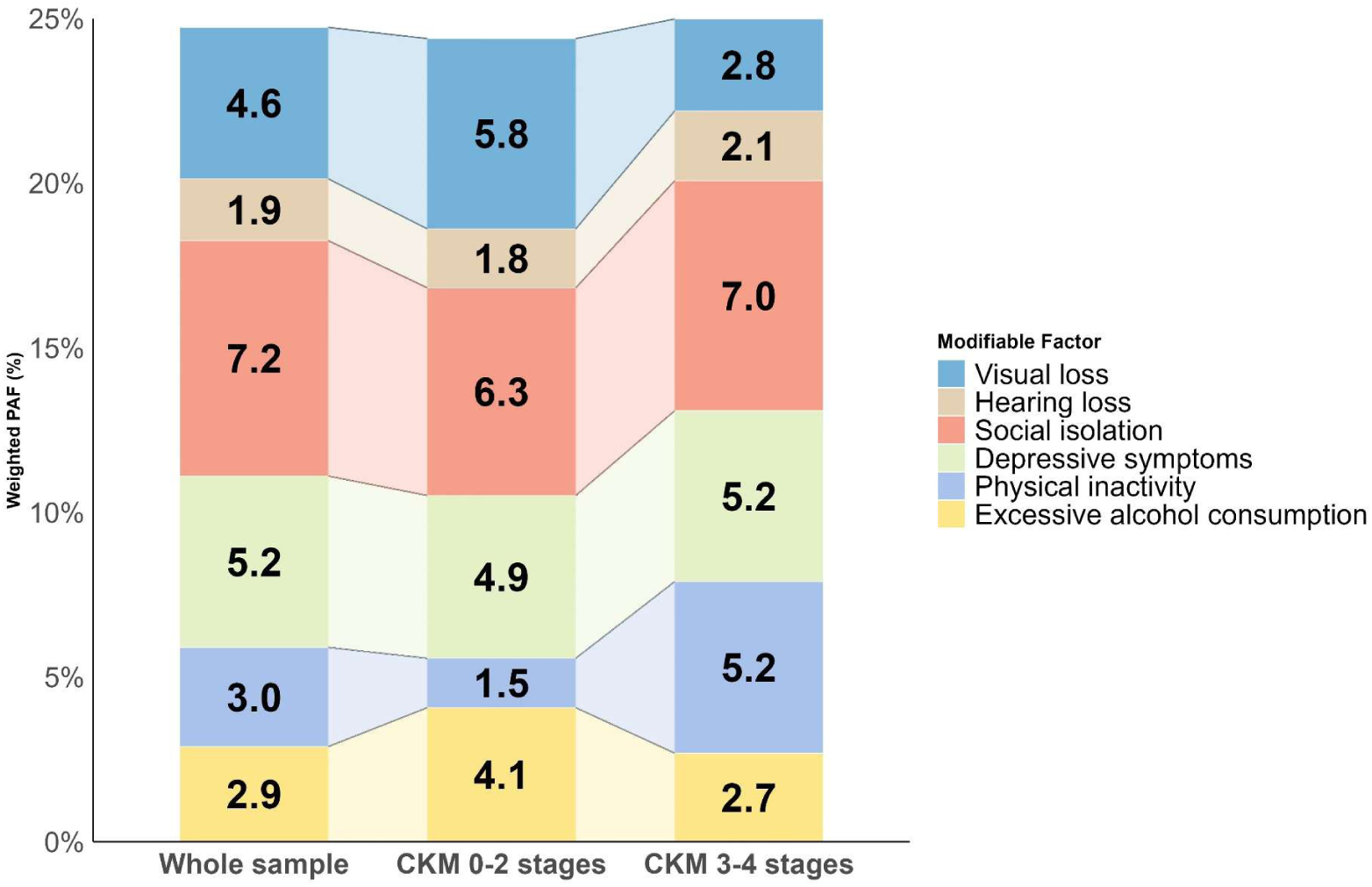
Individual and overall weighted PAF separated by CKM Stages. Note. PAF: Population Attributable Fraction; CKM: Cardiovascular-Kidney-Metabolic Syndrome. Analyses of modifiable factors were conducted exclusively in the global aging cohorts. Weighted individual PAFs were calculated through corresponding hazard ratios, prevalence and, communalities of each modifiable factor. Overall PAF was estimated through the weighted individual PAFs.

### 3.5. Neuroimaging correlates of CKM syndrome

Participant’s characteristics included in the neuroimaging analysis are summarized in Supplemental Table 12. CKM stages were categorized as non-advanced (stages 0-2) and advanced groups (stages 3-4). In cross-sectional analyses between CKM and neuroimaging biomarkers adjusted for age, sex, race, education, SES, and healthy lifestyles, significant differences between the advanced and non-advanced CKM groups were observed for 27 of 34 neuroimaging biomarkers after FDR correction (Supplemental Table 13). The fold changes of the significant biomarkers were ranked and visualized in Figure 3. Notably, the strongest association was observed for white matter hyperintensities (WMHs), with a higher volume associated with the advanced CKM group (fold change = 1.14, p < 0.001). Other key biomarkers associated with advanced CKM stages were reduced accumbens volume (fold change = 0.963 for the left and 0.959 for the right, p < 0.01). Seven biomarkers, including total brain volume, caudate volume, and amygdala volume, did not show significant association with CKM groups.

**Figure 3.**
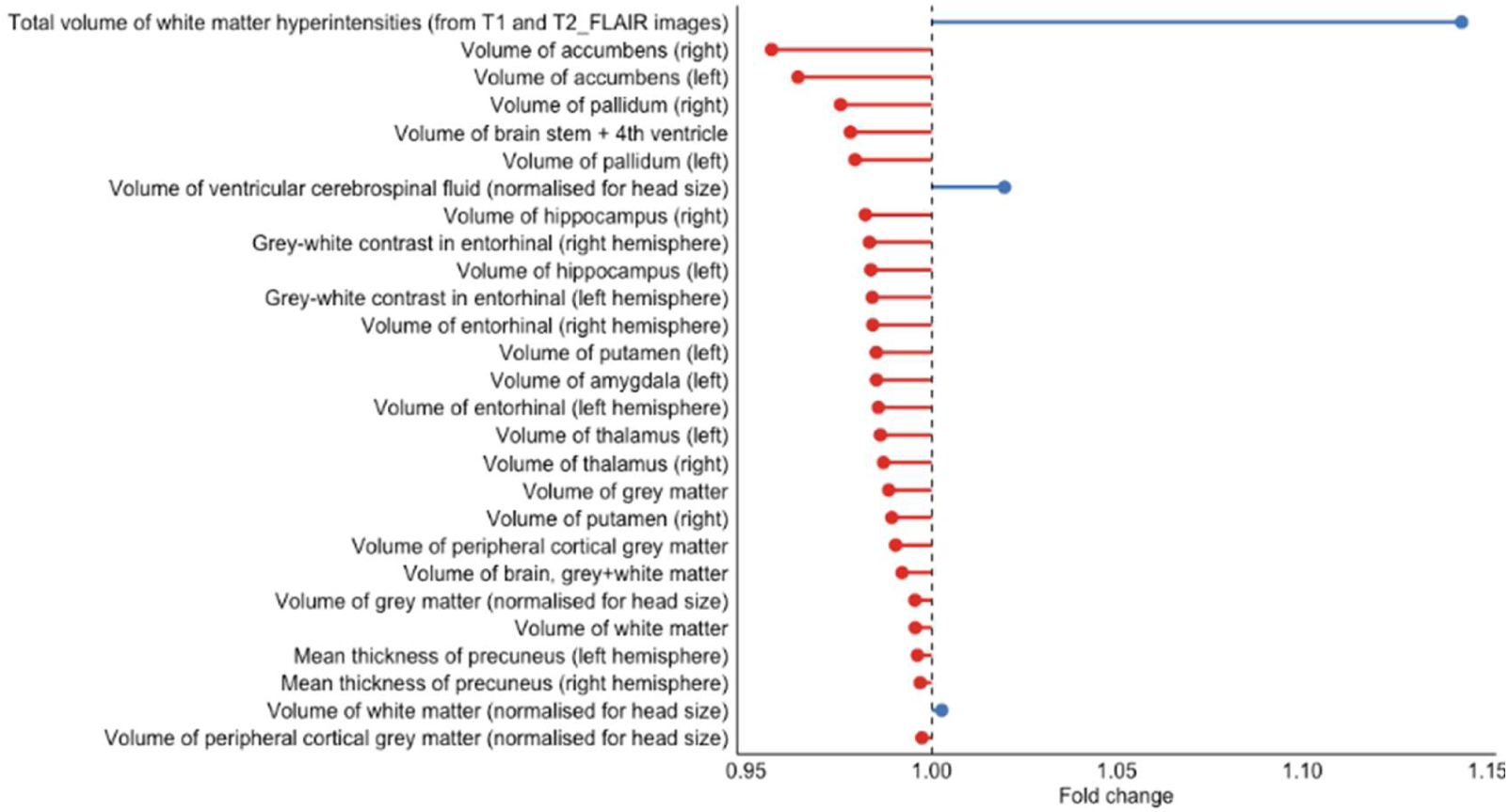
Importance Plot for Brain Imaging Biomarkers associated with Advanced CKM stages. Note. Variable importance ranking of 34 neuroimaging biomarkers from UK Biobank participants (N=29,774), showing their relative associations with advanced CKM stages (3-4) versus non-advanced stages (0-2). Biomarkers are ranked by fold change (x-axis).

### 3.6. Sensitivity analyses

Findings from our main analyses were replicated in sensitivity analyses using multiple imputation with the Markov Chain Monte Carlo method and complete cases (Supplemental Table 14-15). Consistent results were also observed after excluding 188 participants (Supplemental Table 16) who were diagnosed with dementia at the first follow-up after baseline (Supplemental Table 17).

## 4. Discussion

In this large, multi-regional IPD analysis of over 28,242 older adults across 4 continents, with external validation in the international cohorts (COSMIC), we observe a robust stage-dependent relationship between CKM syndrome and cognitive impairment across geographically and culturally diverse cohorts. Extending prior evidence linking severe CKM to dementia risk, our data demonstrates that advanced CKM stages (3–4) confer accelerated global cognitive decline — most prominently in attention, executive function, and language — relative to stages 0–2. Notably, among cognitively intact older adults, advanced CKM stages are further associated with elevated depressive symptomatology. Population-attributable-fraction estimates indicate that modifiable risk factors including physical inactivity, depressive symptoms, social isolation, sensory impairment, and excessive alcohol use together explain 24.8% of incident cognitive-impairment risk within advanced-CKM strata. Moreover, utilizing UKB neuroimaging data, we additionally observe strong coupling between advanced CKM and cerebrovascular pathology, with WMH burden showing the most pronounced association. Taken together, our findings emphasize CKM syndrome as a key multisystem determinant of brain aging and cognitive deterioration and future research should evaluate the implementation of the CKM framework for early identification and prevention of neurocognitive and neuropsychiatric impairment during ageing.

Our findings extend prior evidence^35–37^ by demonstrating associations between CKM syndrome and cognitive decline emerged at stage 3 onward. The CKM framework recognizes the interconnected pathophysiological processes across metabolic, renal, and cardiovascular dysfunctions and provides a staging system that capture their cumulative progression. Importantly, the association between different CKM stages and cognitive function is unlikely to reflect any single component alone. Early CKM stages primarily capture excess adiposity, metabolic risk factors, or mild kidney dysfunction, which may exert relatively subtle effects on the brain. At these stages, compensatory mechanisms, including intact cerebrovascular reserve and neuroplasticity, may buffer against measurable cognitive deterioration.^38^ By contrast, stages 3 and 4 reflect the emergence of subclinical and clinical cardiovascular disease or advanced kidney dysfunction, when cumulative vascular damage, chronic inflammation, and impaired perfusion may increasingly exceed compensatory reserves.^39,40^ This may represent a threshold at which multisystem dysfunction translates into detectable brain changes and cognitive impairment, consistent with the cerebrovascular alterations observed in our neuroimaging analyses. Our findings support prevention strategies aimed at reducing CKM progression before advanced stages 3–4, where the associations with cognitive deterioration become pronounced.

The domain-specific analyses offer insights into potential neuroimaging mechanisms underlying CKM-related cognitive impairment, prominently observed in attention, executive function, and language domains. Reduction in cognitive domains may reflect increased vulnerability to cerebrovascular injury and frontal–subcortical circuit disruption.^41^ Our cross-sectional analysis in the UKB provided neurobiological support for these associations. Advanced CKM stages were associated with brain alterations in 27 of 34 neuroimaging biomarkers, with the strongest effect observed for WMH burden, a well-established marker of cerebral small vessel disease and predictor of cognitive impairment and dementia, especially Alzheimer’s disease.^42^ These findings potentially support a vascular–metabolic hypothesis of cognitive decline, whereby systemic dysfunction leads to cerebral hypoperfusion, and microvascular injury.^43^ Furthermore, volumetric analyses in the UKB revealed modest reductions in cortical volumes and microstructural integrity, including hippocampal alterations, suggesting widespread brain vulnerability. Notably, alterations in the accumbens and other reward-related regions may reflect motivation and reward behaviors, which have been implicated in neuropsychiatric symptoms such as depression and apathy. Future longitudinal studies integrating multimodal biomarkers, including amyloid and tau, are warranted to delineate these mechanisms underlying both neurocognitive and neuropsychiatric disturbances, and identify potentially modifiable points along these pathways.

There are several public health implications of the current findings. First, the findings underscore the importance of early detection and proactive management of CKM progression, particularly before the onset of subclinical and clinical cardiovascular disease. Second, it is essential to implement integrated, multidisciplinary care models that concurrently address metabolic, cardiovascular, renal, and brain health, moving beyond siloed organ-specific approaches. Specifically, consistent with previous studies,^5,6,16^ current findings showed that advanced CKM stages are associated with both neurocognitive and neuropsychiatric disturbances, suggesting potential shared multisystem ageing-related processes. These findings primarily reflect older adults without overt cognitive impairment. Consistent with the study eligibility criteria, excluded participants had a higher prevalence of depressive symptoms, highlighting the need to further examine CKM-related neuropsychiatric features across broader cognitive and psychiatric spectra. Third, we observed that the adverse association of increasing CKM stages on cognitive decline was more pronounced among younger adults, men, and those with lower educational levels, underscoring the need for targeted attention to these vulnerable subgroups.^45^ The presence of APOE ε4 carriage did not significantly modify CKM-cognition associations, suggesting that CKM’s deleterious effect on cognition is independent of established genetic risk factors.^46^ More importantly, associations between modifiable risk factors and incident cognitive impairment highlight the potential targets for risk reduction among individuals with advanced CKM stages. This finding extends previous evidence on the protective effects of non-pharmacological, multidomain lifestyle interventions on cognitive decline to this high-risk population^47^. The six modifiable factors exhibited differing individual weighted PAFs between participants at CKM stages 0–2 and those at advanced stages. Importantly, our results suggest that prioritizing the management of social isolation, depressive symptoms, and physical inactivity may yield greater cognitive benefits among individuals with advanced CKM stages. These factors have been consistently associated with an increased risk of dementia in previous studies^17,48^, and the higher prevalence among individuals with advanced CKM stages may partly explain their greater attributable burden observed in this group.

The current study provides several advantages including the large sample size, long follow-up, and harmonized analyses across multi-regional cohorts, which enhance the generalizability of our findings. The consistency of findings across the COSMIC cohorts, together with convergent neuroimaging findings, provides complementary support for the observed associations. Analyses across diverse populations enhance the generalizability of the findings, while the use of both one-step and two-step IPD approaches helps account for between-study heterogeneity and further supports the robustness of the results. However, several limitations should also be acknowledged. First, harmonizing data across multiple studies inevitably required some loss of granularity and may have introduced misclassification. For example, reliance on self-reported disease diagnoses might lead to underestimation of disease prevalence. Additionally, definitions of CKM components have evolved over time and vary across settings, which may have introduced inconsistencies in diagnosis. Second, CKM stages were assessed at baseline only, precluding evaluation of transitions in stage over time and potentially underestimating the cumulative burden of progression. Future investigation, including repeated CKM measures, is essential to further elucidate the pathways underlying disease progression. Third, residual confounding from unmeasured factors, such as diet or subclinical brain diseases, cannot be excluded. Finally, neuroimaging analyses were cross-sectional and restricted to a subset of relatively healthier UKB participants, limiting causal inference and generalizability to more diverse populations.

To conclude, our findings demonstrate a consistent relationship between CKM syndrome and cognitive decline across multi-regional populations. Advanced CKM stages were associated with accelerated cognitive impairment, particularly in executive, attentional, and language domains, as well as greater depressive symptoms. Given the high prevalence and modifiable nature of CKM, these findings support public health strategies that prioritize early identification and management of CKM risk factors to prevent progression to advanced stages and preserve brain health during ageing worldwide. Among individuals with advanced CKM stages, addressing potentially modifiable factors, particularly social isolation, depressive symptoms, and physical inactivity, may provide additional opportunities for targeted prevention of cognitive impairment.

## Data Availability

For global aging cohorts: The original data for this study are available on their respective websites: the English Longitudinal Study on Ageing (ELSA; https://www.elsa-project.ac.uk), the Survey of Health, Ageing and Retirement in Europe (SHARE; https://share-eric.eu), and the China Health and Retirement Longitudinal Study (CHARLS; http://charls.pku.edu.cn/index/en.html). For COSMIC: Researchers can apply to use COSMIC data by submitting an application form for review by the Research Scientific Committee at Dementias Platform Australia (DPAU): https://portal.dementiasplatform.com.au/data-application.

## Authors’ Contributions

XX, HZ, and LY conceptualized the study. HZ, LY, ZL and WZ performed the data analysis and wrote the first draft. XX and KAT designed the study, developed the protocol and obtained the ethics of this study and conceptualized this study. All authors reviewed the biostatistical results and revised the manuscript, conceived the study and methodology, and interpreted the data. XX and PS were responsible for funding acquisition, data curation, and investigation. All authors revised the manuscript and provided critical revisions to the manuscript. All authors were responsible for reviewing and editing the manuscript.

## Data availability

For global aging cohorts: The original data for this study are available on their respective websites: the English Longitudinal Study on Ageing (ELSA; https://www.elsa-project.ac.uk), the Survey of Health, Ageing and Retirement in Europe (SHARE; https://share-eric.eu), and the China Health and Retirement Longitudinal Study (CHARLS; http://charls.pku.edu.cn/index/en.html).

For COSMIC: Researchers can apply to use COSMIC data by submitting an application form for review by the Research Scientific Committee at Dementias Platform Australia (DPAU): https://portal.dementiasplatform.com.au/data-application.

## Acknowledgments

We thank all team members who have contributed to the ELSA, SHARE, CHARLS studies. This analysis used data or information from the Harmonized ELSA, SHARE, CHARLS datasets, and Codebooks developed by the Gateway to Global Aging Data, supported by the National Institute on Aging (grant numbers R01 AG030153, RC2 AG036619, and 1R03AG043052). For more information, please refer to https://g2aging.org. Funding for ELSA is provided by the National Institute of Aging (grants 2RO1AG7644–01A1 and 2RO1AG017644) and a consortium of UK Government departments coordinated by the Office for National Statistics. SHARE data collection has been funded by European Union (under grant agreement number 101102412) and the European Union’s Horizon 2020 research and innovation program (under grant agreement numbers 870628 and 101015924), and various national funding sources gratefully acknowledged (https://share-eric.eu/infrastructure/funding). CHARLS is supported by the Behavioral and Social Research division of the National Institute on Aging of the National Institute of Health (grants 1-R21-AG031372–01, 1-R01-AG037031–01, and 3-R01AG037031–03S1); the Natural Science Foundation of China (grants 70773002, 0910107022, and 71130002), the World Bank (contracts 7145915 and 7159234), and is conducted by Peking University. We thank all participants for their involvement and the staff of UK Biobank for data collection and participant follow-up.

COSMIC funding from the National Institute On Aging of the National Institutes of Health under Award Number R01AG057531 was awarded to Perminder S. Sachdev, who is also funded by a National Health and Medical Research Council (NHMRC) investigator grant (APP1196150). The head of COSMIC is PSS, and the study coordinator is DML. The research scientific committee leads the scientific agenda of COSMIC and provides ongoing support and governance; it is comprised of member study leaders. The COSMIC research scientific committee and additional principal investigators are listed at https://www.unsw.edu.au/cheba/research-and-impact/consortia/cosmic

The BPRHS was supported by the National Institutes of Health grants P01 AG023394, P50 HL105185, and R01 AG055948. The HELIAD cohort was funded by the Alzheimer’s Association, European Social Fund, and Greek Ministry of Health. The InveCe.Ab study data collection was supported by (wave 1-3) Federazione Alzheimer Italia; Cariplo Foundation (wave 4) and Serpero Foundation (wave 5). The MAS study was funded by the National Health and Medical Research Council (grant numbers APP350833, APP568969, and APP1093083) in Australia.

